# A step forward, but still inadequate: Australian health professionals’ views on the genetics and life insurance moratorium

**DOI:** 10.1101/2021.05.25.21257683

**Authors:** Jane Tiller, Louise Keogh, Aideen McInerney-Leo, Andrea Belcher, Kristine Barlow-Stewart, Tiffany Boughtwood, Penny Gleeson, Grace Dowling, Anya E.R. Prince, Yvonne Bombard, Yann Joly, Martin B Delatycki, Ingrid Winship, Margaret Otlowski, Paul Lacaze

## Abstract

**Background:** In 2019, the Australian life insurance industry introduced a partial moratorium (ban) limiting the use of genetic test results in life insurance underwriting. The moratorium is industry self-regulated and applies only to policies below certain financial limits (eg AUD$500,000 of life cover).

**Methods:** We surveyed Australian health professionals (HPs) who discuss genetic testing with patients, to assess knowledge of the moratorium; reported patient experiences since its commencement; and HP views regarding regulation of genetic discrimination (GD) in Australia.

**Results:** Between April-June 2020, 166 eligible HPs responded to the online survey. Of these, 86% were aware of the moratorium, but <50% had attended related training/information sessions. Only 16% answered all knowledge questions correctly, yet 69% believed they had sufficient knowledge to advise patients. Genetics HPs’ awareness and knowledge were better than non-genetics HPs’ (p<0.05). There was some reported decrease in patients delaying/declining testing after the moratorium’s introduction, however 42% of HPs disagreed that patients were more willing to have testing post-moratorium. Although many (76%) felt the moratorium resolved some GD concerns, most (88%) still have concerns, primarily around self-regulation, financial limits and the moratorium’s temporary nature. Almost half (49%) of HPs reported being dissatisfied with the moratorium as a solution to GD. The majority (95%) felt government oversight is required, and 93% felt specific Australian legislation regarding GD is required.

**Conclusion:** While the current Australian moratorium is considered a step forward, most HPs believe it falls short of an adequate long-term regulatory solution to GD in life insurance.

## Introduction

Genetic discrimination (GD) is an area of international concern[1-4]. In the context of life insurance, GD can lead to increased premiums or denial of insurance applications. Predictive genetic testing (where an individual has not developed disease but genetic testing can reveal a higher risk of developing disease) can save lives due to preventative interventions or early treatment of disease. In Australia and internationally, research shows that fear of insurance implications deters some at-risk individuals from having clinically-indicated predictive genetic testing or participating in research[5-10].

In Australia, the issue of GD in health insurance does not arise, because health insurance premiums are community-rated rather than personally risk-rated under the *Private Health Insurance Act* 2007 (Cth). However, life insurance companies can legally ask for and use applicant’s genetic test results to make underwriting decisions, under a specific exemption in s46 of the *Disability Discrimination Act* 1992 (Cth) (DDA). Despite consideration of the potential for GD arising from this exemption in 2001-03[11], the Australian government continues to allow the life insurance industry to self-regulate their access to and use of applicants’ genetic information. The inherently conflicted model of industry self-regulation for life insurance raises numerous ethical and societal concerns[12]. These have been reflected in government inquiries into the insurance and financial services industries in recent years[13 14].

In 2018, the Australian Parliamentary Joint Committee on Corporations and Financial Services released recommendations arising out of its inquiry into the life insurance industry[14]. These included recommending a ban (moratorium) on life insurers’ use of predictive genetic test results for underwriting, as well as recommending government monitoring and introduction of legislation if necessary. The Australian government has not yet responded to these recommendations. However, in 2019, the Financial Services Council (FSC), the peak industry body for Australian life insurers, voluntarily introduced an industry self-regulated moratorium on the use of genetic test results by their member organisations[15 16]. The FSC moratorium, which applies only to FSC member companies, restricts insurers’ access to and use of genetic test results for applications for life insurance cover for or below AUD$500,000 only. Because travel insurance falls within general insurance, as distinct from life insurance, it is not restricted by the moratorium and genetic test results can legally be used to discriminate in underwriting, pursuant to s46 of the DDA.

As a self-regulated industry standard, the FSC moratorium is not legally binding or enforceable; nor is it subject to government regulation or oversight. Legally, the insurance industry’s right to use genetic test results in underwriting in accordance with the DDA remains and is not affected by the implementation of the FSC moratorium, which will be reviewed in 2022 and end in 2024, if not renewed. The FSC have a Code of Practice, compliance with which is monitored by an external committee of three persons including a consumer representative, an industry representative and an independent chair[17]. At the time of publication, the FSC moratorium has not been incorporated into the Code of Practice, although we understand that this is FSC’s intention in future.

Health professionals (HPs) are key to ensuring that patients who are considering genetic testing are adequately advised of the potential for life insurance discrimination before testing is undertaken. Further, they often witness the deterrent effects of fears about life insurance implications, or hear firsthand of GD experiences[18]. Australian professional guidelines state that a discussion of insurance implications should be part of each genetic counselling session where relevant[19]. With the progressive mainstreaming of genetic testing in Australia, a greater proportion of clinicians without genetics training are discussing genetic testing with patients[20]. A recent systematic review[21] found non-genetics HPs (nurses and physicians) had limited genetics knowledge and were unprepared for integrating genomics into clinical care. However, little is known about non-genetics HPs’ knowledge regarding life insurance discrimination, the moratorium, and the implications for patients.

Prior to the introduction of the FSC moratorium, we surveyed genetics professionals in Australia about their patients’ experiences regarding life insurance discrimination and their views on regulation of this area [18]. That study demonstrated some deficits both in knowledge regarding use of genetic testing in insurance underwriting, and self-reported confidence in advising patients about insurance implications. It also captured widespread concerns regarding regulation in the area of GD, with the vast majority of HPs reporting a view that current Australian regulations were inadequate to protect patients from GD.

To our knowledge, there has been no survey of HPs following the introduction of the FSC moratorium. This study forms a key part of the Australian Genetics and Life Insurance Moratorium: Monitoring the Effectiveness and Response (A-GLIMMER) Project - funded by the Australian government[22 23]. The purpose of the Project is to assess the extent to which the FSC moratorium achieves the policy aims identified by the Parliamentary Joint Committee on Corporations and Financial Services. This particular study contributes to that Project by analysing the effectiveness of the FSC moratorium in relation to HPs. The aim of this study is to describe the knowledge, experiences and perspectives of HPs that discuss genetic testing with patients, following the commencement of the FSC moratorium. Where possible, it will also compare those findings with findings from the pre-moratorium research study[18].

## Methods

### Population, sampling and recruitment

The protocol paper for the A-GLIMMER project has been published previously[23]. The population of interest was qualified health professionals working in a health service in Australia, who regularly discuss genetic testing with patients. Eligibility was established through screening questions at the beginning of the questionnaire. A range of targeted recruitment strategies were adopted to capture a broad sample:

- Newsletters emailed directly to members of the Human Genetics Society of Australasia (HGSA), Australasian Society of Genetic Counsellors, Royal Australasian College of Physicians, and the Australian Genomics Health Alliance
- Social media advertisements (Twitter and Facebook)
- Direct email to colleagues and personal contacts of the authors
- Snowball sampling (requesting HPs and personal contacts forward an email invitation to their professional networks)

### Survey development and data collection

We used an online survey conducted using REDCap software[24] (see Supplementary Materials for a copy of the survey). The survey was adapted from our previous survey of HPs administered prior to the commencement of the FSC moratorium[18]. Relevant questions were preserved for comparison, and new questions were introduced to determine the impact and effectiveness of the moratorium in relation to HPs’ knowledge, experience and views. The adapted survey included sections relating to demographics; awareness, training, and knowledge; patient attitudes, behaviours, and reported experiences; and personal views regarding regulation of GD. Validated scales were unavailable for questions specific to the moratorium, however the survey was developed in consultation with a number of clinical and research partners, and was pre-tested with a clinical geneticist, a genetic counsellor and a lay person without health qualifications. Data were mostly collected through closed-ended responses using Likert scales and fixed alternative options, with a small number of open-ended questions where free text was allowed. The survey was open for response from April–June 2020,

### Data analysis

Descriptive analysis was conducted for closed-ended questions. Descriptive statistics were reported for each question included in the results, broken down by total number of HPs, as well as separately by genetics HPs and non-genetics HPs. Six questions evaluated knowledge (true/false/unsure) about aspects of the FSC moratorium and current insurance implications). HPs received a point for every correct answer (range 0–6). A mean knowledge score was calculated for comparison between groups. The percentage of correct answers for each item was also calculated. Knowledge scores were categorised into “good knowledge” (5-6 questions answered correctly), “average knowledge” (3-4 correct), and “poor knowledge” (0-2 correct). Z-tests were used to test for significance of differences between groups, with p-values (2-sided) <0.05 considered significant. STATA 14 was used for analysis[25].

Responses to open-ended questions were reviewed and sorted into common categories, which were reported in detail in the supplementary materials and in summary form with example quotes in the manuscript.

## Results

Overall, 166 eligible HPs participated in the online survey. As some HPs discontinued the survey before reaching the end, the number of HPs who answered each individual question varied (range n=144-166). To aid readability, the “n” for every reported figure is not given in the text, but is included at each instance in the accompanying figures and tables. Given the diverse recruitment strategies, it is difficult to estimate a total response rate relative to all eligible participants. However, at the time of recruitment, the HGSA distribution list included 484 clinical geneticists (CGs) and genetic counsellors (GCs). Of the 166 HPs who participated, 111 were CGs or GCs, making the response rate for those categories in the profession field an estimated 23%.

Table 1 shows the characteristics of the surveyed population. The “Other” category under the profession field is comprised of HPs representing more than 15 different fields, all of whom were eligible for the study as they reported regularly discussing genetic testing with patients. These included surgery, nursing, psychiatry, metabolics, cardiology, pathology, neurology, endocrinology, gastroenterology, haematology, immunology, obstetrics, paediatrics, and others (see Supplementary Table S1). These HPs are referred to as “non-genetics HPs”, as distinguished from “genetics HPs” (genetic counsellors, clinical geneticists, and genetics fellows).

**Table 1:**
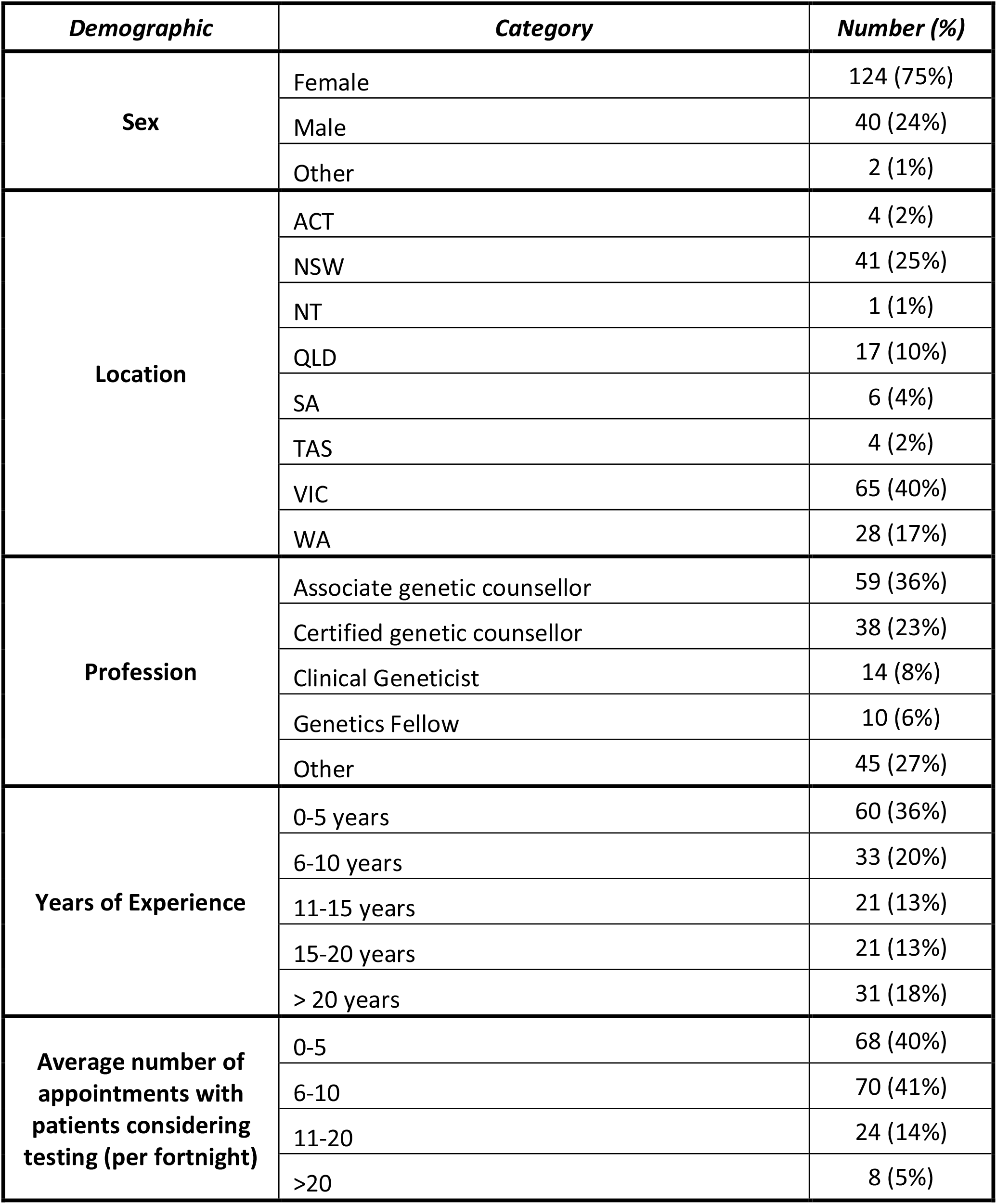
Characteristics of the surveyed population (n=166)

| ***Demographic*** | ***Category*** | ***Number (%)*** |
| --- | --- | --- |
| **Sex** | Female | 124 (75%) |
|  | Male | 40 (24%) |
|  | Other | 2 (1%) |
| **Location** | ACT | 4 (2%) |
|  | NSW | 41 (25%) |
|  | NT | 1 (1%) |
|  | QLD | 17 (10%) |
|  | SA | 6 (4%) |
|  | TAS | 4 (2%) |
|  | VIC | 65 (40%) |
|  | WA | 28 (17%) |
| **Profession** | Associate genetic counsellor | 59 (36%) |
|  | Certified genetic counsellor | 38 (23%) |
|  | Clinical Geneticist | 14 (8%) |
|  | Genetics Fellow | 10 (6%) |
|  | Other | 45 (27%) |
| **Years of Experience** | 0-5 years | 60 (36%) |
|  | 6-10 years | 33 (20%) |
|  | 11-15 years | 21 (13%) |
|  | 15-20 years | 21 (13%) |
|  | > 20 years | 31 (18%) |
| **Average number of appointments with patients considering testing (per fortnight)** | 0-5 | 68 (40%) |
|  | 6-10 | 70 (41%) |
|  | 11-20 | 24 (14%) |
|  | >20 | 8 (5%) |

### Awareness, knowledge and training (Table 2 and Figure 1)

Most HPs (86%) overall, but just over half (53%) of non-genetics HPs were aware of the FSC moratorium. Over half of genetics HPs (55%) reported attending training or information sessions regarding the moratorium and insurance implications of genetic testing training, while few non-genetics HPs had done so (7%). There are two well-known fact sheets on the moratorium: the Centre for Genetics Education (CGE) Fact Sheet 20[26], and the FSC insurance and genetics moratorium fact sheet[27]. A majority of HPs (65%) had read at least one of these fact sheets. However, only a third (n=14) of non-genetics HPs had read a fact-sheet, compared to 76% (n=89) of genetics HPs (z=5; p<0.05).

**Table 2:** Awareness, knowledge, training.

| Question |  | Responses | Genetics HPs (%) | Non-genetics HPs (%) | TOTAL (%) |
| --- | --- | --- | --- | --- | --- |
| Are you aware that there was a change in policy on 1 July 2019 and a moratorium was introduced on the use of genetic testing in life insurance underwriting? (n=166) |  | No | 3/121 (2) | 21/45 (47) | 24/166 (14) |
|  |  | Yes | 118/121 (98) | 24/45 (53) | 142/166 (86) |
| [if yes] How did you become aware? (n=142)<br>* more than one option could be selected |  | My health service | 64/118 (54) | 7/24 (29) | 71/142 (50) |
|  |  | A news source | 12/118 (10) | 10/24 (42) | 22/142 (15) |
|  |  | HGSA | 96/118 (81) | 4/24 (17) | 100/142 (70) |
|  |  | Insurance industry | 4/118 (3) | 0/24 (0) | 4/142 (3) |
| Has your health service provided, or have you attended, any training or information sessions regarding the moratorium and insurance implications of genetic testing? (n=166) |  | Yes, formal training | 7/121 (6) | 0/45 (0) | 7/166 (4) |
|  |  | Yes, information sessions | 60/121 (49) | 3/45 (7) | 63/166 (38) |
|  |  | No | 54/121 (45) | 42/45 (93) | 96/166 (58) |
| How well do you feel you now understand insurance implications for individuals undergoing genetic testing? (n=166) |  | Extremely well | 12/121 (10) | 0/45 (0) | 12/166 (7) |
|  |  | Reasonably well | 89/121 (74) | 17/45 (38) | 106/166 (64) |
|  |  | Not particularly well | 17/121 (14) | 17/45 (38) | 34/166 (20) |
|  |  | Not well at all | 3/121 (2) | 11/45 (24) | 14/166 (8) |
| Do you feel you have sufficient knowledge about the current insurance implications of genetic testing to properly advise patients? (n=166) |  | Yes | 98/121 (81) | 16/45 (36) | 114/166 (69) |
|  |  | No | 23/121 (19) | 29/45 (64) | 52/166 (31) |
| Are you aware of, and have you read, these fact sheets? (n=158) | The updated HGSA position statement on Genetic Testing and Life Insurance (updated after announcement of moratorium) | I am aware of it and I have read it | 49/117 (42) | 9/42 (21) | 58/158 (37) |
|  |  | I am aware of it, but have not yet read it | 42/117 (36) | 13/42 (31) | 55/158 (35) |
|  |  | I am not aware of it | 26/117 (22) | 20/42 (48) | 46/158 (29) |
|  | Fact Sheet 20 published by the Centre for Genetics Education (updated mid-2019) | I am aware of it and I have read it | 79/117 (68) | 7/42 (17) | 86/158 (54) |
|  |  | I am aware of it, but have not yet read it | 17/117 (15) | 6/42 (14) | 23/158 (15) |
|  |  | I am not aware of it | 21/117 (18) | 28/42 (67) | 49/158 (31) |
|  | The Financial Services Council (FSC) Standard No 11 on Genetic testing (updated to include the moratorium in mid-2019) | I am aware of it and I have read it | 29/117 (25) | 6/42 (14) | 35/158 (22) |
|  |  | I am aware of it, but have not yet read it | 42/117 (36) | 7/42 (17) | 49/158 (31) |
|  |  | I am not aware of it | 46/117 (39) | 28/42 (67) | 74/158 (47) |
|  | The FSC fact sheet on the life insurance moratorium | I am aware of it and I have read it | 51/117 (44) | 11/42 (26) | 62/158 (39) |
|  |  | I am aware of it, but have not yet read it | 18/117 (15) | 2/42 (5) | 20/158 (13) |
|  |  | I am not aware of it | 48/117 (41) | 28/42 (67) | 76/158 (48) |
| Number of knowledge questions answered correctly (n=146)<br>(for question-specific data see Supplementary Table S2)<br>Mean score (Genetics HPs): 4.5<br>Mean score (Non-genetics HPs): 3.1 | 0 | "Poor knowledge" | 1/110 (1) | 4/36 (11) | 5/146 (3) |
|  | 1 |  | 0/110 (0) | 5/36 (14) | 5/146 (3) |
|  | 2 |  | 1/110 (1) | 4/36 (11) | 5/146 (3) |
|  | 3 | "Average knowledge" | 14/110 (13) | 7/36 (19) | 21/146 (14) |
|  | 4 |  | 34/110 (31) | 5/36 (14) | 39/146 (27) |
|  | 5 | "Good knowledge" | 41/110 (37) | 7/36 (19) | 48/146 (33) |
|  | 6 |  | 19/110 (17) | 4/36 (11) | 23/146 (16) |

**Figure 1:**
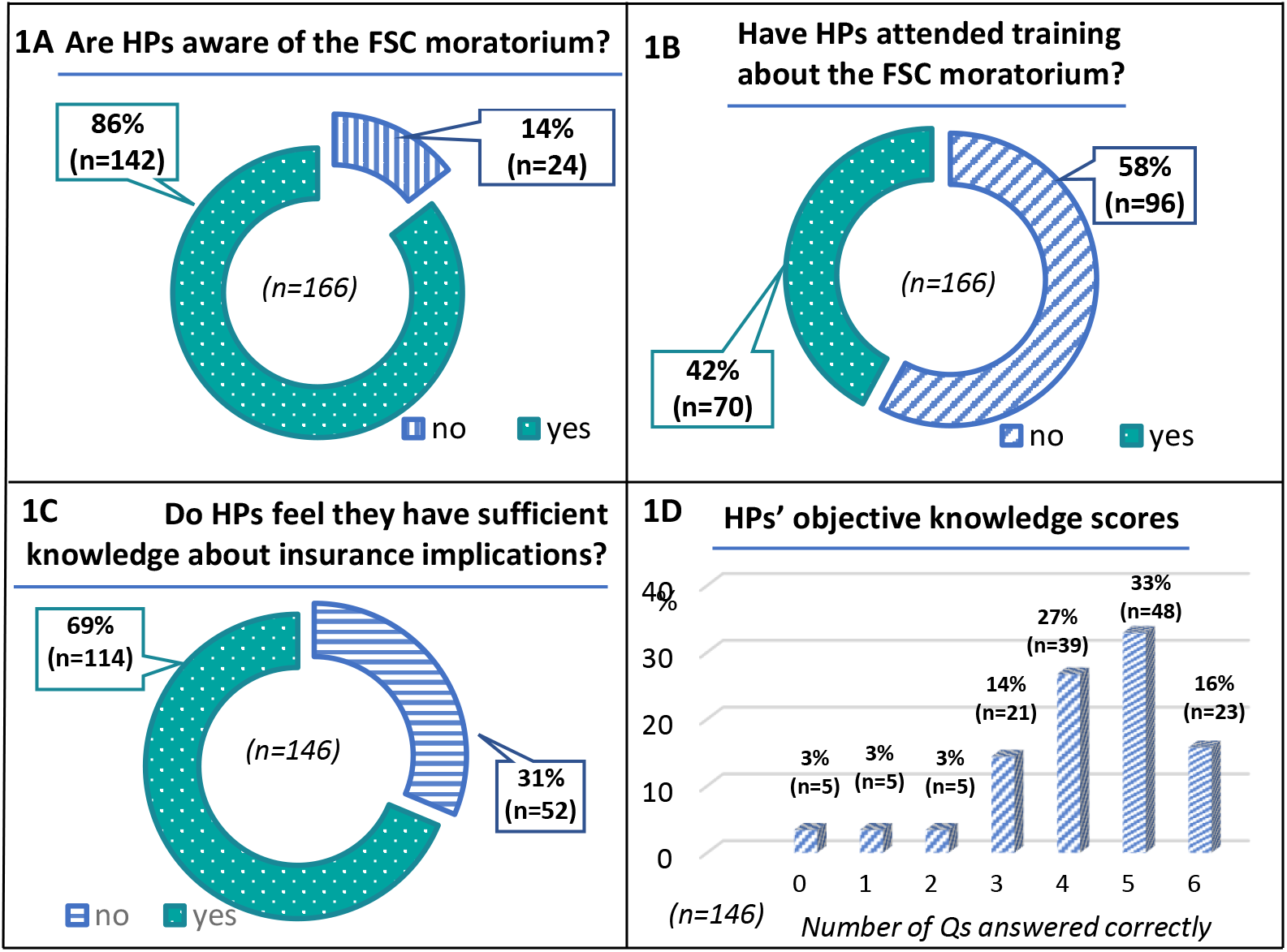
Awareness, knowledge, training

Many HPs (69%) felt that they had sufficient knowledge about current insurance implications to properly advise clients. On the objective knowledge test, about half (49%) had good knowledge (5-6 questions answered correctly) (see Table S2 for question-specific data). More genetics HPs answered questions correctly (mean 4.5/6) than non-genetics HPs (mean 3.1/6) (z=7.3; P<0.05). Of the 50 genetics HPs who answered two or more questions incorrectly (average/poor knowledge), about three-quarters (n=38) felt they understood insurance implications for individuals undergoing genetic testing extremely/reasonably well, and almost two-thirds (n=30) felt they had sufficient knowledge to properly advise patients. However, of 25 non-genetics HPs who answered two/more incorrectly, only a quarter felt they understood insurance implications extremely/reasonably well, (n=6) and a fifth (n=5) felt they had sufficient knowledge. These differences between genetics and non-genetics HPs were significant (z=4.3 (understanding) z=3.3 (knowledge), p<0.05).

### Impact on practice and testing (Table 3)

HPs were asked in this survey about how often patients either delayed or declined having predictive genetic testing due to life insurance concerns, both before the moratorium was introduced and then after the moratorium was introduced (Fig 2A and 2B). Overall, 63% of HPs said patients delayed testing because of life insurance concerns often/sometimes before the moratorium, and 39% said they delayed often/sometimes after the moratorium was introduced (z=4.15; p<0.05). Similarly, 39% said patients refused testing due to life insurance concerns often/sometimes pre-moratorium, compared with 18% post-moratorium (z=4.18; p<0.05).

**Table 3:** Impact on practice and clients.

| Question |  | Responses | Genetics HPs (%) | Non-genetics HPs (%) | TOTAL (%) |
| --- | --- | --- | --- | --- | --- |
| Is there a statement about insurance implications.. (n=150) | On your consent form, where you have a specific form for predictive genetic testing in adults (n=51) | Yes | 34/38 (89) | 9/13 (69) | 43/51 (84) |
|  |  | No | 4/38 (11) | 4/13 (31) | 8/51 (16) |
|  | On your consent form, where you have a standard form for all genetic testing (n=99) | Yes | 60/75 (80) | 6/24 (25) | 66/99 (67) |
|  |  | No | 15/75 (20) | 18/24 (75) | 33/99 (33) |
| Has your consent form been updated following the introduction of the moratorium on 1 July 2019? (n=151) |  | Yes | 24/113 (21) | 4/38 (11) | 28/151 (18) |
|  |  | No | 67/113 (59) | 8/38 (21) | 75/154 (50) |
|  |  | I don't know | 22/113 (19) | 26/38 (68) | 48/154 (32) |
| How often do you estimate patients delayed predictive testing (n=154) | Due to life, income or trauma/critical illness insurance concerns, before the moratorium was introduced? | Never | 1/121 (1) | 5/45 (11) | 6/154 (4) |
|  |  | Rarely | 41/121 (34) | 11/45 (24) | 52/154 (34) |
|  |  | Sometimes | 68/121 (56) | 19/45 (42) | 87/154 (56) |
|  |  | Often | 5/121 (4) | 5/45 (11) | 10/154 (6) |
|  | Due to life, income or trauma/critical illness insurance concerns, after the moratorium was introduced? | Never | 12/121 (10) | 5/45 (11) | 17/154 (11) |
|  |  | Rarely | 62/121 (51) | 15/45 (33) | 77/154 (50) |
|  |  | Sometimes | 40/121 (33) | 18/45 (40) | 58/154 (38) |
|  |  | Often | 0/121 (0) | 2/45 (4) | 2/154 (1) |
|  | Due to travel insurance concerns? | Never | 47 /121 (39) | 14/45 (31) | 61/154 (40) |
|  |  | Rarely | 60/121 (49) | 16/45 (36) | 76/154 (49) |
|  |  | Sometimes | 7/121 (6) | 5/45 (11) | 12/154 (8) |
|  |  | Often | 0/121 (0) | 5/45 (11) | 5/154 (3) |
| How often do you estimate patients refused predictive testing (n=154) | Due to life, income or trauma/critical illness insurance concerns, before the moratorium was introduced? | Never | 11/121 (9) | 7(16) | 17/154 (11) |
|  |  | Rarely | 76/121 (63) | 12/45 (27) | 77/154 (50) |
|  |  | Sometimes | 26/121 (21) | 19/45 (42) | 58/154 (38) |
|  |  | Often | 2 /121 (2) | 2/45 (4) | 2/154 (1) |
|  | Due to life, income or trauma/critical illness insurance concerns, after the moratorium was introduced? | Never | 28/121 (23) | 6/45 (13) | 34/154 (22) |
|  |  | Rarely | 75/121 (62) | 18/45 (40) | 93/154 (60) |
|  |  | Sometimes | 11/121 (9) | 15/45 (33) | 26/154 (17) |
|  |  | Often | 0/121 (0) | 1/45 (2) | 1/154 (1) |
|  | Due to travel insurance concerns? | Never | 56/121 (46) | 16/45 (36) | 72/154 (47) |
|  |  | Rarely | 50/121 (41) | 14/45 (31) | 64/154 (42) |
|  |  | Sometimes | 8/121 (7) | 8/45 (18) | 16/154 (10) |
|  |  | Often | 0/121 (0) | 2/45 (4) | 2/154 (1) |
| Since the introduction of the moratorium, have patient/s told you about having had an adverse insurance outcome on the basis of genetic test results? (for example, having difficulty obtaining a policy, having an increased premium, or having a policy application denied)? (n=153) |  | Yes | 3/114 (3) | 2/39 (5) | 5/153 (3) |
|  |  | No | 111/114 (97) | 37/39 (95) | 148/153 (97) |

**Figure 2:**
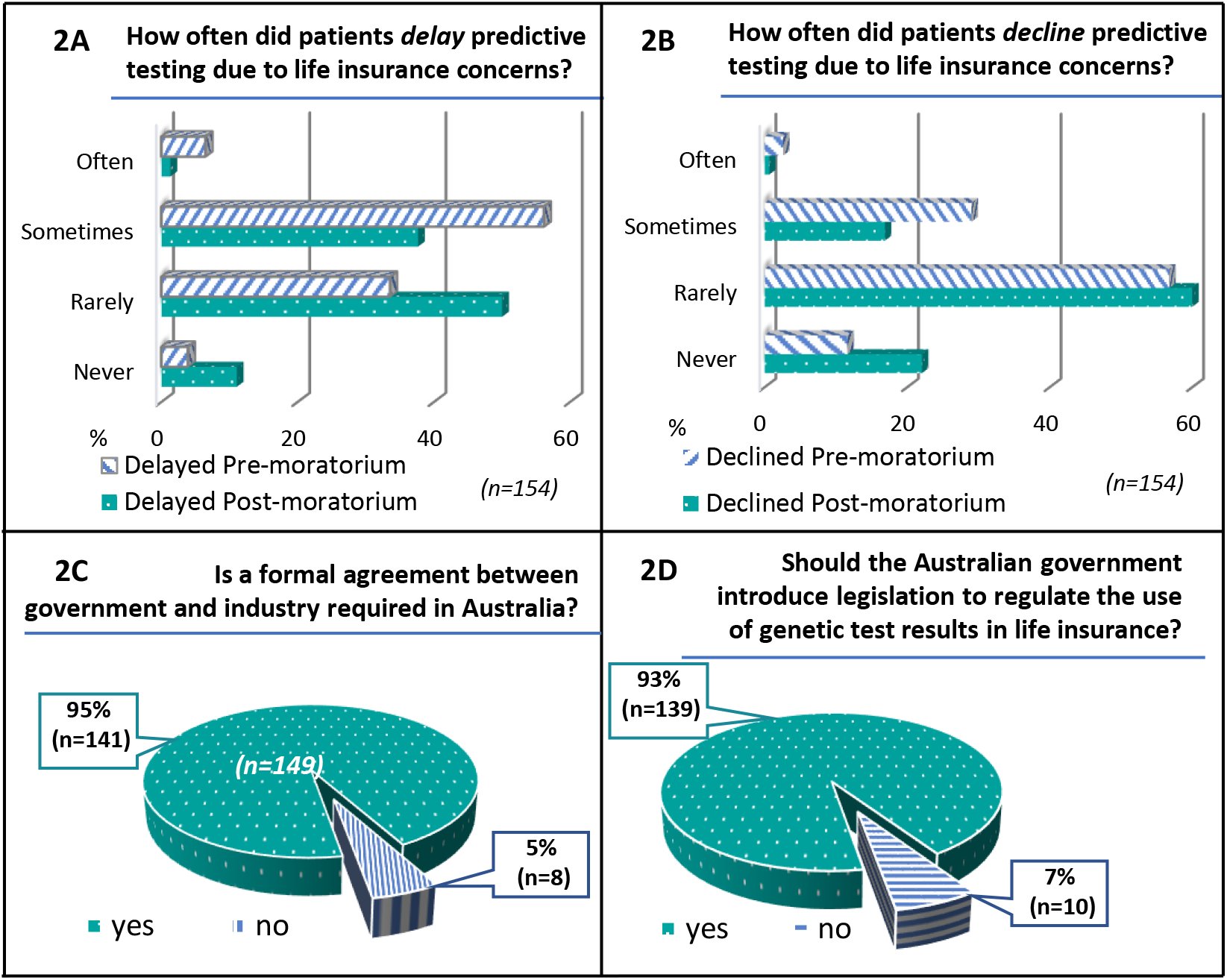
Patient impact and views on regulation

Although the FSC moratorium does not apply to travel insurance, this can be a source of some confusion for both patients and consumers. GD in travel insurance was raised as an issue by several HPs in free-text responses (see below). When asked about how often patients delay or decline predictive testing due to travel insurance concerns, 11% of HPs said patients delay often/sometimes, and 12% said patients refuse often/sometimes.

### Views on the FSC moratorium’s effectiveness and regulation (Table 4)

Almost all HPs (93%) agreed/strongly agreed that “*consumers are better protected post-moratorium than they were before the moratorium was introduced*”. Although 76% felt the FSC moratorium resolved some concerns the HPs had about insurance discrimination, 88% still had concerns about insurance discrimination after its introduction.

**Table 4:** Views on FSC moratorium and regulation.

| Question |  | Responses | Genetics HPs (%) | Non-genetics HPs (%) | TOTAL (%) |
| --- | --- | --- | --- | --- | --- |
| Please indicate the degree to which you agree with the following statements | The moratorium is easy to understand (n=145) | Strongly agree | 0/113 (0) | 0/35 (0) | 0/145 (0) |
|  |  | Agree | 89/110 (81) | 27/35 (77) | 116/145 (80) |
|  |  | Disagree | 18/110 (16) | 6/35 (17) | 24/145 (17) |
|  |  | Strongly disagree | 3/110 (3) | 2/35 (6) | 5/145 (3) |
|  | The moratorium is easy to explain to patients (n=144) | Strongly agree | 1/110 (1) | 0/35 (0) | 1/144 (1) |
|  |  | Agree | 80/109 (73) | 28/35 (80) | 108/144 (75) |
|  |  | Disagree | 25/109 (23) | 5/35 (14) | 30/144 (21) |
|  |  | Strongly disagree | 3/109 (3) | 2/35 (6) | 5/144 (3) |
|  | Patients are less confused than they used to be about insurance implications of genetic testing (n=144) | Strongly agree | 5/109 (5) | 0/35 (0) | 5/144 (3) |
|  |  | Agree | 51/109 (47) | 17/35 (49) | 68/144 (47) |
|  |  | Disagree | 49/109 (45) | 17/35 (49) | 66/144 (46) |
|  |  | Strongly disagree | 4/109 (4) | 1/35 (3) | 5/144 (3) |
|  | Patients are more willing to have predictive genetic testing than they were before the moratorium was introduced (n=144) | Strongly agree | 6/109 (5) | 0/35 (0) | 6/144 (4) |
|  |  | Agree | 60/109 (55) | 19/35 (54) | 79/144 (55) |
|  |  | Disagree | 39/109 (36) | 15/35 (43) | 54/144 (38) |
|  |  | Strongly disagree | 4/109 (4) | 1/35 (3) | 5/144 (3) |
|  | The moratorium has resolved some concerns I had about insurance discrimination (n=144) | Strongly agree | 5/109 (5) | 2/35 (6) | 7/144 (5) |
|  |  | Agree | 82/109 (75) | 21/35 (60) | 103/144 (72) |
|  |  | Disagree | 21/109 (19) | 10/35 (29) | 31/144 (22) |
|  |  | Strongly disagree | 1/109 (1) | 2/35 (6) | 3/144 (2) |
|  | After the introduction of the moratorium, I still have concerns about insurance discrimination (n=144) | Strongly agree | 22/109 (20) | 4/35 (11) | 26/144 (18) |
|  |  | Agree | 76/109 (70) | 25/35 (71) | 101/144 (70) |
|  |  | Disagree | 9/109 (8) | 4/35 (11) | 13/144 (9) |
|  |  | Strongly disagree | 2/109 (2) | 2/35 (6) | 4/144 (3) |
|  | Consumers are better protected post-moratorium than they were before the moratorium was introduced (n=144) | Strongly agree | 18/109 (16) | 2/35 (6) | 20/144 (14) |
|  |  | Agree | 84/109 (77) | 30/35 (86) | 114/144 (79) |
|  |  | Disagree | 7/109 (6) | 3/35 (9) | 10/144 (7) |
|  |  | Strongly disagree | 0/109 (0) | 0/35 (0) | 0/144 (0) |
| Based on your professional experience, how do you feel about the moratorium as a solution to genetic discrimination in life insurance? (n=149) |  | Very satisfied | 6/113 (5) | 0/36 (0) | 6 /149 (4) |
|  |  | Somewhat satisfied | 52/113 (46) | 18/36 (13) | 70/149 (47) |
|  |  | Somewhat dissatisfied | 49/113 (46) | 12/36 (9) | 61/149 (41) |
|  |  | Very dissatisfied | 6 /113 (5) | 6/36 (4) | 12/149 (8) |
| In your opinion, how should insurers' compliance with the moratorium on using genetic test results in life insurance be regulated? (n=149)<br>* more than one option could be selected |  | Self-regulation by the life insurance industry (FSC) | 16/113 (14) | 4/36 (11) | 20/149 (13) |
|  |  | Regulation through legally enforceable rules | 102/113 (90) | 29/36 (81) | 131/149 (88) |
|  |  | Other | 2/113 (2) | 3/36 (8) | 5/149 (3) |
| In the UK, there is a moratorium that involves a formal agreement between the UK government and the Life Insurance Industry. Do you think a formal agreement between the Australian government and industry (Financial Services Council) is required on this issue in Australia? (n=149) |  | Yes | 108/113 (96) | 33/36 (92) | 141/149 (95) |
|  |  | No | 5/113 (4) | 3/36 (8) | 8/149 (5) |
| Do you think the Australian government should introduce legislation to regulate the use of genetic test results in life insurance? (n=149) |  | Yes | 105/113 (93) | 34/36 (94) | 139/149 (93) |
|  |  | No | 8/113 (7) | 2/36 (6) | 10/149 (7) |

Most HPs agreed/strongly agreed that the FSC moratorium is easy to understand (80%) and easy to explain to patients (76%); however, a number (20% and 24% respectively), disagreed/strongly disagreed, showing a portion of HPs find it difficult to understand and/or explain. HPs were split almost evenly in their views regarding both questions “*patients are less confused than they used to be about insurance implications of genetic testing*” (51% agreed/strongly agreed; 49% disagreed/strongly disagreed) and “*patients are more willing to have predictive genetic testing than they were before the moratorium was introduced*” (59% agreed/strongly agreed; 41% disagreed/strongly disagreed).

The vast majority (95%) of HPs (with no significant difference between genetics and non-genetics HPs (z=0.2; p=0.83)) stated that a formal agreement between government and the life insurance industry (as exists in the UK) is required in Australia (Fig 2C). This question allowed optional free text to allow for HPs to elaborate on their answer (see Table S3 for all free-text responses grouped into categories). Of 149 HPs, 22 elected to elaborate (21 who said yes, 1 who said no). Of those who said yes, one-third expressed concerns with industry self-regulation. For example, Participant 129 (genetic counsellor, 0-5 years’ experience) stated, “*I think that the industry needs to be held accountable; I don’t trust that the self-governing model is enough*.*”* Two HPs felt that further regulation may be needed, but the decision should depend on the outcomes of the FSC moratorium, with Participant 127 (genetic pathologist, 15-20 years’ experience) stating, “*We need an evidence-based approach. We should wait for results to emerge from the current moratorium*.” The HP who said no and chose to elaborate (Participant 109, clinical geneticist, 15-20 years’ experience) stated, “*Insurance companies currently load premiums or withhold cover on much less scientific premises than genetic test results. By making these ‘special’ we do more harm than good by making people afraid of genetic testing and complicating the process”*.

The vast majority (93%) of HPs also indicated that the Australian government should introduce legislation to regulate the use of genetic test results in life insurance (no significant difference between genetics/non-genetics HPs (z=-0.1; p=0.94)) (Fig 2D). Of 149 HPs, 15 elaborated (13 “yes”; 3 “no”) (Table S3). Four HPs expressed mistrust of insurers, with Participant 207 (genetic counsellor, >20 years’ experience) stating, “*if it is not in law, why would an insurance company do it?”*. Four HPs commented that legally-enforceable/legislation-based regulation is required to ensure consumer protection; for example, Participant 135 (clinical geneticist, >20 years’ experience) noted, “*this is the only way to protect people properly and not have the highly undesirable situation where people don’t have genetic testing because of insurance concerns and die of preventable disease*.*”*

One HP’s reason (Participant 256, Registered Nurse, >20 years’ experience) for answering “no” to the government regulating insurer use of genetic information through legislation, appeared to be that insurer use should not be allowed *at all*, stating, “*Sorry, too many instances where insurance companies look to preserving their cash and not interested in helping people with genuine need*.” Participant 229 (clinical geneticist, 15-20 years’ experience) answered no “*with the caveat that self-regulation is effective and sufficient monitoring is in place*” along with 2 others who stated that any regulation should be evidence-based. The other “no” HP (Participant 109, clinical geneticist, 15-20 years’ experience) explained that, “*People accept that information available will be used by insurance companies. They don’t generally have a problem with this*”.

When asked about how insurers’ compliance with the FSC moratorium on using genetic test results in life insurance should be regulated, 88% of HPs chose “regulation through legally enforceable rules”. Thirteen percent (n=20) chose self-regulation by the FSC, though 7 of these also chose “legally enforceable rules” indicating a preference for a blended regulatory approach. Forty-nine percent of HPs felt either very or somewhat dissatisfied with the moratorium as a solution to GD in life insurance. Only 4% felt “very satisfied”.

### Benefits and limitations of the moratorium (Table 5)

Sixty-two HPs responded to the optional free-text question “In your opinion, what, if any, are/have been the benefits of the moratorium?” Table 5a sets out the categories of benefits that were expressed, with example quotes (see Supplementary Table S4 for full responses). The most common responses were “increased reassurance/patients more willing to have genetic testing” (34%) and “some protection provided” (31%). Sixty-one HPs provided optional feedback to the question, “in your opinion, what, if any, are the limitations of the moratorium?” (Table 5b, Table S4). The most commonly raised limitations were “insurance companies’ compliance/ self-regulation” (46%), “financial limits” (44%), and “temporary nature of the moratorium” (31%). Similar issues arose in free-text comments in the final question, “Do you have any final comments?” (Table S5). Of 21 HPs who responded with substantive comments about the FSC moratorium, a third (n=7) raised issues around the need for legislation/enforceability; 2 each expressed concerns with the temporary nature and the unjustness of discrimination based on uncontrollable factors; 1 reiterated the inadequacy of the financial limits; 5 reported difficulty with understanding/explaining the moratorium to patients; and 3 expressed concerns with the applicability to travel insurance. No HPs made any positive comments about the moratorium in the final thoughts section.

**Table 5a:** Perceived *benefits* of the current genetics and life insurance moratorium in Australia.

| Benefit (n=62) | n (%)<sup>*</sup> | Example quote(s) | Participant #<br>(qualification, yrs' experience) |
| --- | --- | --- | --- |
| Increased reassurance/<br>patients more willing to have genetic testing | 21 (34%) | *Easing concerns for patients who may now have some level of cover if at high genetic risk. By doing this it lessens the potential negative implications of predictive testing and therefore decision making can be focused more on the health implications.* | P21 (GC, 10-15 y) |
|  |  | *It's a step in the right direction and patients with minor concerns/reluctance feel reassured* | P108 (GC, 0-5 y) |
| Some protection provided | 19 (31%) | *Provides at least some level of insurance that may not have been available at all previously* | P42 (GC, 10-15 y) |
|  |  | *People can access some level of insurance without the threat of discrimination based on their genetic test result* | P199 (GC, 0-5 y) |
| Increased clarity | 9 (15%) | *From my practice point of view, having some clear guidelines to present to clients/patients, rather than it all being very dependent on the individual insurer.* | P129 (GC, 0-5 y) |
| Family implications | 6 (10%) | *Most people are concerned about what the insurance implications are for their children. It is helpful to be able to let them know that their children only need to disclose their parent's health conditions not their genetic test result.* | P136 (GC, 0-5 y) |
| Heightened awareness/<br>recognition of issue | 5 (8%) | *More awareness of the issue, hopefully future stronger protections for patients depending on how effective the moratorium can be shown to be currently.* | P130 (GC, 6-10 y) |
| "Step in the right direction" | 3 (5%) | *It is a step in the right direction but insurance concerns are still present for many patients and providers* | P108 (GC, 0-5 y) |
| Provides time | 2 (3%) | *Gives time to find better solution* | P98 (GC, 15-20 y) |
| * participants may have listed multiple benefits in their free-text response<br>GC=Genetic Counsellor; CG = Clinical Geneticist |  |  |  |

**Table 5b:** Perceived *limitations* of the current genetics and life insurance moratorium in Australia.

| Limitation<br>(n=64) | n<br>(%)* | Example quote(s) | Participant #<br>(qualification, yrs' experience) |
| --- | --- | --- | --- |
| Insurance companies' compliance/ self-regulation | 29<br>(45%) | *It would be better if there was NO discrimination at all, that was made law and insurance companies held accountable (not self-regulated).* | P129 (GC, 0-5 y) |
|  |  | *It is self-regulated and not legally enforceable, so only as good as the trust in the industry generally* | P89 (GC, 0-5 y) |
| Financial limits | 28<br>(44%) | *The limit on cover is relatively low. Despite industry assurance that most policies fall below this threshold a significant number of patients see this as limiting* | P229 (CG 15-20 y) |
|  |  | *The amounts are too low and won't give enough reassurance to some.* | P135 (CG >20 y) |
| Temporary nature of moratorium | 17<br>(27%) | *The uncertainty about how long it will be in place - we need this to be PERMANENT to enable patients not to fear having genetic testing because of insurance concerns as genetic testing can really influence their physical AND psychological health* | P149 (GC 15-20 y) |
|  |  | *The uncertainty of how this will apply in the future if someone wants to take out a policy in a few years and the moratorium no longer applies.* | P173 (GC, 0-5 y) |
| Restricted application | 8<br>(13%) | *Not all insurers are FSC Members. It doesn't apply to all life insurance policies, only those under certain amounts. Only applies to policies from 1 July 2019, ie not pre-existing too.* | P42 (GC, 11-15 y) |
| Travel insurance not covered | 3<br>(5%) | *Travel insurance is a major exclusion. Many patients are concerned about implications for travel insurance especially when their work or family takes them to high cost medical care in countries such as USA.* | P195 (GC, 15-20 y) |
| Lack of dissemination | 2<br>(3%) | *Many financial advisors and workers in the industry seem unaware of the moratorium.* | P207 (CG, >20 y) |
| * participants may have listed multiple limitations in their free-text response<br>GC=Genetic Counsellor; CG = Clinical Geneticist |  |  |  |

## Discussion

In this study, we surveyed Australian health professionals’ (HPs) knowledge, experiences, and opinions regarding the current industry self-regulated moratorium on genetic testing and life insurance. We found that most HPs who discuss genetic testing with clients are aware of the FSC moratorium, though knowledge of key aspects of the moratorium could be improved. Both awareness of and knowledge about the moratorium are superior for genetics HPs than non-genetics HPs. Many HPs expressed a view that the moratorium had resolved some of their concerns with GD. However, the majority of HPs still have concerns regarding the potential for GD in life insurance and feel that the moratorium does not adequately address those concerns. Most HPs feel that the moratorium does not represent an adequate long-term regulatory solution for Australia. Specifically, the majority of HPs feel that more stringent consumer protections are required, especially in the form of stronger government regulation or legislation. Key findings of our study are summarized in Figure 3.

**Figure 3:**
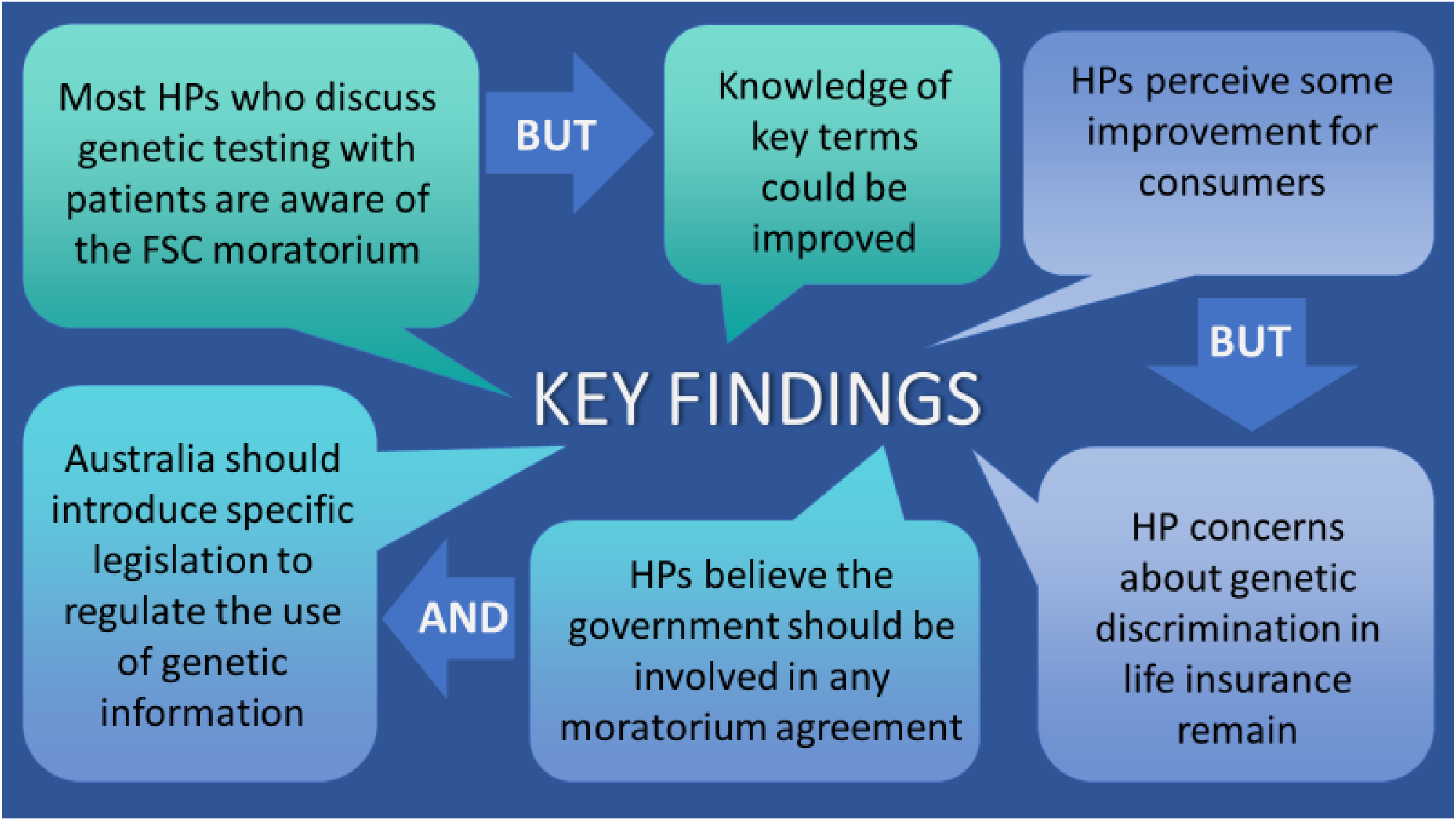
Key findings

In our previous survey of Australian genetics HPs, conducted before the FSC moratorium was introduced[18], we found that only 9% (n=6/69) of HPs felt regulation at the time was adequate. Now, after the introduction of the FSC moratorium, we still find that >90% believe government regulation and legislation are required. Although the moratorium is seen as a step forward, most Australian HPs remain concerned about the potential for GD, and its impact on patients. We found evidence of recognition from HPs regarding improved consumer protections, compared with the pre-moratorium situation, with most HPs agreeing that consumers are better protected now, after the introduction of the FSC moratorium. When asked about benefits of the moratorium, some HPs cited increased willingness of patients to have genetic testing. However, despite some reported reduction in patients delaying or refusing testing for insurance reasons, more than 40% of HPs still disagreed that patients are more willing to have testing following the introduction of the FSC moratorium, suggesting the desired impact of the moratorium has not been fully achieved.

Although about half of HPs surveyed expressed some satisfaction with the FSC moratorium as a solution to GD in life insurance, about half were either somewhat or very dissatisfied with it, and a vast majority still had concerns about insurance discrimination post-moratorium. Primary concerns, expressed in free-text comments, centred around the nature of industry self-regulation, lack of government oversight, the financial limits on the moratorium and its temporary nature. Comments provided by HPs showed negative opinions - including distrust of insurers, the conflicted nature of industry self-regulation, the need for more stringent government regulation, the inadequacy of the current financial limits, and the temporary nature of the FSC moratorium (and the uncertainty this creates for patients in the future). A small minority of HPs felt that government regulation was not required, as either the moratorium was adequate or the government should wait and see whether it is effective before introducing further regulation.

Many countries have banned or restricted life insurers’ access to genetic test results for underwriting purposes[28-30]. For example, Canada has implemented the *Genetic Nondiscrimination Act* (2017) (GNDA), which prevents insurers from using genetic test results, and the US’ *Genetic Information Nondiscrimination Act* (2008) (GINA) bans the use of genetic test results in health insurance and employment contexts. The UK’s moratorium (known since 2018 as the Code on Genetic Testing and Insurance[31]) was established in 2001 as an agreement between the insurance industry and the UK Government, to protect consumers in relation to the use of predictive genetic test results. A single exception applies to predictive genetic tests for Huntington’s disease where the life insurance policy coverage is above £500,000 (∼AUD$910,000).

Almost all HPs surveyed believe that a formal agreement between government and industry is required for Australia. Further, most HPs felt that any moratorium should be regulated through legally enforceable rules from the Australian government, including specific legislation to regulate life insurers’ use of genetic test results. Our findings demonstrate that HPs who offer genetic testing to patients believe that the current policy situation is still inadequate and lacks sufficient consumer protections. Given that in our previous study 62% of HPs considered Australia should introduce such legislation, and 93% of current HPs considered that legislation was needed, it appears that the current FSC moratorium has not altered that perception for the majority of HPs.

Although the FSC moratorium may soon be included in the FSC Code of Practice[17], compliance with this Code is monitored by a committee of three individuals and is not subject to any legal or regulatory oversight from government. There are no pathways for enforcement by consumers (other than complaints to the Financial Ombudsman Service), and the sanctions which can be imposed lack any legal weight or punitive power. Thus, inclusion of the moratorium in the Code in future may not alleviate HPs concerns regarding lack of government oversight of this issue.

The demographics of HPs in this study are similar to those of our previous study[18]. However, the current cohort is larger (n=166, compared with n=87 previously) and more diverse due to the expanded recruitment strategy. This survey has highlighted the diversity of health professionals who are discussing genetic testing with patients, in line with the mainstreaming of genetic testing noted earlier. This has also captured, for the first time in Australia, the perceptions of these HPs on this issue.

Although there was consensus among most HPs from genetics and non-genetics backgrounds on key issues, including regulation, there was also divergence between genetics and non-genetic HPs in some areas. Areas of divergence include awareness of the FSC moratorium, with only about half of non-genetics HPs who discuss genetic testing with patients being aware of the moratorium. Our results in this regard are consistent with other studies that report poor awareness or understanding of local non-discrimination laws/policies by HPs [32-34]. However, given the importance of considering insurance issues where relevant before deciding whether to have a genetic test, this lack of awareness is somewhat concerning, and raises questions about dissemination and how to more effectively raise awareness, particularly among non-genetics HPs. The numbers of genetics and genetics HPs who had read either the CGE or FSC fact sheets indicate that these are a reasonable method of disseminating information to genetics HPs but not so effective for non-genetics HPs.

Another area of distinction between genetics and non-genetics HPs was in objective knowledge. When only considering genetic HPs, 81% felt they had sufficient knowledge of insurance implications to properly advise patients, which has increased from our previous research (61%; n=53/87)[18]. However, only a small fraction of both genetics and non-genetics HPs answered all 6 questions about key aspects of the FSC moratorium correctly, and about half had average or poor knowledge. For non-genetics HPs, there was a reasonable match between subjective and objective lack of knowledge. This is consistent with international studies of non-genetics HPs, which found a correlation between HPs’ subjective and objective knowledge levels regarding genetic non-discrimination regulations[35] and genetics generally [36]. However, although genetics HPs were more knowledgeable than non-genetics HPs, they appeared to overestimate their knowledge more than non-genetics HPs, indicating some mismatch between subjective and objective knowledge.

An area of historical misinformation is that of the impact of GD on health insurance. In our previous survey[18], 15% of HPs incorrectly stated that genetic information could be used for health insurance policies in Australia. In the current survey, a similar number (17%) of genetics HPs still answered this question incorrectly, as well as 50% of non-genetics HPs. The knowledge gap between genetic and non-genetic HPs overall was sizable, highlighting the need to train a wider range of HPs with the mainstreaming of genetic testing. Surprisingly, given the recent policy changes and need for dissemination and education around these changes, similar numbers of genetics HPs reported attending training in our previous survey (51%) as this survey (55%). Further, a smaller percentage of genetics HPs reported having read the CGE fact sheet (68%) than previously reported (85%)[18]. This may explain the knowledge gaps despite clinician confidence (HPs who feel they have sufficient knowledge may be less likely to seek out additional resources).

A significant finding of the study is that many HPs (50%) believe the FSC moratorium applies to travel insurance, or are unsure of its application. As discussed, travel insurers are not restricted by the moratorium. Several HPs raised concerns about insurance implications for travel insurance in free-text comments regarding limitations of the moratorium and in the “final thoughts” section. This provides further support for the contention that broader government regulation and oversight of the use of genetic test results in insurance underwriting is required to adequately protect consumers.

Strengths of the current study include being the first of its kind to report HP views and experiences since the introduction of the FSC moratorium. Also, to our knowledge, our study provides the first example of a survey of HPs conducted both before and after the introduction of a major policy change regarding GD and life insurance. By asking similar questions as our previous (pre-moratorium) survey, we were able to undertake comparative analysis pre- and post-moratorium. Our survey reached a wide range of HPs, covering traditional genetics HPs as well as other clinicians who discuss genetic testing with patients.

Limitations of our study include the relatively small number of non-genetics HPs surveyed, which may limit the generalisability to this group. HPs were asked some questions about patient experiences, which is arguably second-hand information. Other studies, which will seek firsthand experiences/perceptions of patients and consumers, are being developed as part of the A-GLIMMER Project[37] to address this limitation. Given the rising prominence of the issue of life insurance discrimination in Australia, response bias is a potential limitation. We attempted to address this by making it an option for HPs to remain anonymous if they preferred. Further, views expressed by HPs who were happy to be contacted (∼20% of HPs) will be explored further through qualitative interviews, in a subsequent study. Our survey was conducted less than a year after the introduction of the FSC moratorium (9 months). Although this was intentional to ensure data collection and analysis could take place to inform the review of the moratorium, waiting longer could have resulted in different responses and experiences from HPs. As the survey was conducted online and in early 2020, it is not expected that COVID restrictions significantly affected participation.

## Conclusion

Many Australian genetic HPs know about the FSC moratorium and have knowledge of its specifics; however, some genetic HPs and many non-genetics HPs do not. Australian HPs report some improvement for consumers as a result of the moratorium’s introduction, but concerns about GD in life insurance remain. HPs describe strong views about perceived limitations of the moratorium, including industry self-regulation and lack of government oversight, as well as the inadequacy of the current financial limits and the uncertainty around the moratorium’s temporary nature. A majority of Australian HPs believe government oversight of the FSC moratorium is required, and that legislation regarding genetic testing and life insurance should also be considered in Australia. Our findings will assist with developing recommendations for the Australian government to consider future policy and regulatory changes in this area, and will be of interest to other jurisdictions internationally who are grappling with similar issues around the regulation of GD in life insurance.

## Supporting information

Supplementary Materials - online survey

## Data Availability

Numerous data are made available via supplementary materials. Additional data can be made available on reasonable request.

## Declarations

### Funding (information that explains whether and by whom the research was supported)

The project is supported by a grant from the Australian Government’s Medical Research Future Fund (MRFF), ref 76721. AML is funded by a National Health and Medical Research Council (NHMRC) Early Career Fellowship (ID 1158111). PL is supported by a National Heart Foundation Future Leader Fellowship (ID 102604).

### Conflicts of interest/Competing interests (include appropriate disclosures)

The authors declare no conflicts of interest or competing interests

### Code availability (software application or custom code)

N/A

## Authors’ contributions (optional: please review the submission guidelines from the journal whether statements are mandatory)

### Ethics approval (include appropriate approvals or waivers)

This project was granted approval by the Monash University Human Research Ethics Committee on 11 March 2020, ID number 22576, and was performed in accordance with the ethical standards as laid down in the 1964 Declaration of Helsinki.

### Consent to participate (include appropriate statements)

The Explanatory Statement provided to participants includes the following statement: “Participation is completely voluntary and you are able to withdraw at any time. However, if you chose to remain anonymous in completing the survey, we will not be able to identify your data in order to exclude it from analysis.”

The first question of the online survey completed by participants (following the reading of the Explanatory Statement) was as follows:

“By continuing with this survey, you give your consent to being a participant in this research project

- Continue with survey
- I do not want to continue with this survey” (survey ended if this response was selected)

### Consent for publication (include appropriate statements)

The Explanatory Statement provided to participants includes the following statement: “The data collected will be de-identified so that the source of each response is not known, and the de-identified data will be analysed and submitted for publication in a peer-reviewed journal.”

## Notes

### Competing Interest Statement

The authors have declared no competing interest.

### Clinical Protocols

https://rdcu.be/ck58P

