## Supplementary Materials - online survey for "A step forward, but still inadequate: Australian health professionals’ views on the genetics and life insurance moratorium"

### Genetics and insurance: Post-moratorium survey of health professionals

#### Explanatory Statement and consent

Please download and read the Explanatory Statement and indicate your willingness to proceed with the survey

[Attachment: "Explanatory statement health professionals.pdf"]

| | |
|---|---|
| By continuing with this survey, you give your consent to being a participant in this research project | ◯ Continue with survey<br>◯ I do not want to continue with this survey |
| Are you a qualified health professional (other than a general practitioner*)? | ◯ Yes<br>◯ No<br>(* Please note that the scope of this survey does not extend to GPs) |
| Do you work in a setting in which you have direct contact with patients who are considering genetic testing? | ◯ Yes<br>◯ No |
| Do you work in either Australia or New Zealand? | ◯ Australia<br>◯ New Zealand<br>◯ No |
| Because the situation is slightly different in NZ than Australia, we have set up a customised survey just for you. Please click on this box and this survey will close and take you to the NZ survey. Thanks for participating! :) | ◯ Continue |
| Thank you for your willingness to participate in the survey. Unfortunately, you do not meet the criteria for inclusion. Thanks anyway! | ◯ close survey |

#### DEMOGRAPHIC INFORMATION

| | |
|---|---|
| Sex | ◯ Male<br>◯ Female<br>◯ Other/prefer not to say |
| Profession | ◯ Clinical geneticist<br>◯ Genetics fellow<br>◯ Associate genetic counsellor<br>◯ Certified genetic counsellor<br>◯ Oncologist<br>◯ Genetic pathologist<br>◯ Other |
| Profession: | __________________________________ |
| I have been practising as a [profqual] for | ◯ 0-5 years<br>◯ 6-10 years<br>◯ 11-15 years<br>◯ 15-20 years<br>◯ more than 20 years |
| I have been practising as an [profqual] for | ◯ 0-5 years<br>◯ 6-10 years<br>◯ 11-15 years<br>◯ 15-20 years<br>◯ more than 20 years |
| I have been practising as a [profother] for | ◯ 0-5 years<br>◯ 6-10 years<br>◯ 11-15 years<br>◯ 15-20 years<br>◯ more than 20 years |
| On average, the number of formal appointments I take with patients who are considering genetic testing, by phone or in person, per fortnight, is | ◯ 0-5<br>◯ 6-10<br>◯ 11-20<br>◯ more than 20 |
| The health service where I primarily work is located in | ◯ ACT<br>◯ NSW<br>◯ NT<br>◯ QLD<br>◯ SA<br>◯ TAS<br>◯ VIC<br>◯ WA |
| The health service where I primarily work is in the | ◯ Private sector<br>◯ Public sector<br>◯ Both |
| The health service where I primarily work is | ◯ Urban<br>◯ Regional/rural |

| | |
|---|---|
| I see/speak with patients who are considering testing in the following scenarios [tick all that apply] | ☐ Diagnostic testing in adults<br>☐ Diagnostic testing in children<br>☐ Predictive testing in unaffected adults<br>☐ Predictive testing in unaffected children<br>☐ Carrier testing for recessive conditions in adults<br>☐ Prenatal testing<br>☐ The return of secondary findings from clinical testing<br>☐ The return of secondary findings from research<br>☐ Other |
| Other: | __________________________________ |

#### TRAINING AND EDUCATION

Did you participate in the previous survey on genetics and insurance in 2017?

- ◯ Yes
- ◯ No
- ◯ I do not remember

Are you aware that there was a change in policy on 1 July 2019 and a moratorium was introduced on the use of genetic testing in life insurance underwriting?

- ◯ Yes
- ◯ No

How did you become aware? [select all that apply]

- ☐ Through my health service
- ☐ Through a news source or social media
- ☐ Through the HGSA or another professional body
- ☐ Through the insurance industry directly
- ☐ Other

Other: __________________________________

Has your health service provided, or have you attended, any training or information sessions regarding the moratorium and insurance implications of genetic testing? [select all that apply]

- ☐ Yes, formal training
- ☐ Yes, information sessions
- ☐ No

Do you feel this training has been adequate?

- ◯ Yes
- ◯ No

How well do you feel you now understand insurance implications for individuals undergoing genetic testing?

- ◯ Extremely well
- ◯ Reasonably well
- ◯ Not particularly well
- ◯ Not well at all

Do you feel you have sufficient knowledge about the current insurance implications of genetic testing to properly advise patients?

- ◯ Yes
- ◯ No

#### PROFESSIONAL PRACTICE

The moratorium is a self-regulated (regulated by the insurance industry, not by government) policy change. From 1 July 2019, life insurers have agreed not to ask for or use applicants' genetic test results when underwriting policies worth up to

- $500,000 for life cover,
- $200,000 for trauma/critical illness cover, and
- $4000/month for income protection.

For policies worth over this amount, life insurers will still be able to use genetic test results when underwriting.

**With regard to each of the following fact sheets regarding insurance and genetics, please indicate whether you are aware of and whether you have read each.**

**You can access and view copies of these documents below if you wish.**

| | I am aware of it and I have read it | I am aware of it, but have not yet read it | I am not aware of it |
|---|---|---|---|
| The updated HGSA position statement on Genetic Testing and Life Insurance (updated after announcement of moratorium) | ○ | ○ | ○ |
| Fact Sheet 20 published by the Centre for Genetics Education (updated mid-2019) | ○ | ○ | ○ |
| The Financial Services Council (FSC) Standard No 11 on Genetic testing (updated to include the moratorium in mid-2019) | ○ | ○ | ○ |
| The FSC fact sheet on the life insurance moratorium | ○ | ○ | ○ |

[Attachment: "HGSA updated position statement - Genetic Testing and Life Insurance in Australia.pdf"]

[Attachment: "Centre for Genetics Education Fact sheet 20 - LIFE INSURANCE PRODUCTS AND GENETIC TESTING IN AUSTRALIA.pdf"]

[Attachment: "FSC Standard 11 - Moratorium.pdf"]

[Attachment: "FSC fact sheet - Moratorium Key Facts (26 June 2019).pdf"]

**Before the introduction of the moratorium, how often do you estimate patients delayed or refused predictive genetic testing because of  life, income or trauma/critical illness insurance concerns?**

| | Often | Sometimes | Rarely | Never |
|---|---|---|---|---|
| Delayed | ○ | ○ | ○ | ○ |
| Refused | ○ | ○ | ○ | ○ |

**Following the introduction of the moratorium, how often do you estimate patients delay or refuse predictive genetic testing because of  life, income or trauma/critical illness insurance concerns?**

| | Often | Sometimes | Rarely | Never |
|---|---|---|---|---|
| Delay | ○ | ○ | ○ | ○ |
| Refuse | ○ | ○ | ○ | ○ |

**How often do you estimate patients delay or refuse genetic testing because of travel insurance concerns?**

| | Often | Sometimes | Rarely | Never |
|---|---|---|---|---|
| Delay | ◯ | ◯ | ◯ | ◯ |
| Refuse | ◯ | ◯ | ◯ | ◯ |

Are you, or have you been, involved in recruiting participants into research studies?

◯ Yes
◯ No

Before the introduction of the moratorium, how often do you estimate participants refused or were concerned about being involved with genetic RESEARCH because of life, income or disability insurance concerns?

| | Often | Sometimes | Rarely | Never |
|---|---|---|---|---|
| Refused | ◯ | ◯ | ◯ | ◯ |
| Concerned | ◯ | ◯ | ◯ | ◯ |

Following the introduction of the moratorium how often do you estimate participants refuse or are concerned about being involved with genetic RESEARCH because of life, income or disability insurance concerns?

| | Often | Sometimes | Rarely | Never |
|---|---|---|---|---|
| Refuse | ◯ | ◯ | ◯ | ◯ |
| Concerned | ◯ | ◯ | ◯ | ◯ |

Since the introduction of the moratorium, have patient/s told you about having had an adverse insurance outcome on the basis of genetic test results? (for example, having difficulty obtaining a policy, having an increased premium, or having a policy application denied)?

◯ Yes
◯ No

Please provide further details (if applicable):

__________________________________________

Does your health service now have an agreed policy regarding communicating with patients about insurance implications of genetic testing?

◯ Yes, a written policy
◯ Yes - a verbal policy that has been discussed with me or at meetings at which I was present
◯ No
◯ I don't know

Does this policy include a specific reference to the moratorium?

◯ Yes
◯ No
◯ I don't know

**I discuss insurance implications with clients in the following scenarios:**
**(only those which you previously selected will appear here)**

| | always | sometimes | never |
|---|---|---|---|
| Diagnostic testing in adults | ○ | ○ | ○ |
| Diagnostic testing in children | ○ | ○ | ○ |
| Predictive testing in unaffected adults | ○ | ○ | ○ |
| Predictive testing in unaffected children | ○ | ○ | ○ |
| Carrier testing for recessive conditions in adults | ○ | ○ | ○ |
| Prenatal testing | ○ | ○ | ○ |
| The return of secondary findings from clinical testing | ○ | ○ | ○ |
| The return of secondary findings from research | ○ | ○ | ○ |
| [testtypeother] | ○ | ○ | ○ |

You indicated that you [predadult:checked] discuss insurance implications for predictive testing in adults. Why is this? __________________________________

#### CONSENT

| | |
|---|---|
| When obtaining consent for genetic testing, does your health service have a specific form for predictive testing in adults? | ◯ Yes<br>◯ No, there is one standard consent form used for all testing |
| Does the standard consent form include a statement about insurance implications? | ◯ Yes<br>◯ No |
| Does the form contain a statement about insurance implications? | ◯ Yes<br>◯ No |
| Has your consent form been updated following the introduction of the moratorium on 1 July 2019? | ◯ Yes<br>◯ No<br>◯ I don't know |
| Further details (if applicable): | __________________________________________ |

#### PERSONAL VIEWS

We understand that you are a health professional and not a legal or insurance professional. However, we are interested in your personal views on the following matters, based on your experience as a health professional.

The moratorium is a self-regulated (regulated by the insurance industry, not by government) policy change. From 1 July 2019, life insurers have agreed not to ask for or use applicants' genetic test results when underwriting policies worth up to

- $500,000 for life cover,
- $200,000 for trauma/critical illness cover, and
- $4000/month for income protection.

For policies worth over this amount, life insurers will still be able to use genetic test results when underwriting.

Based on your professional experience, how do you feel about the moratorium as a solution to genetic discrimination in life insurance?

◯ Very satisfied - this is the ideal solution
◯ Somewhat satisfied - this is a pretty good solution
◯ Somewhat dissatisfied - the solution could be better
◯ Very dissatisfied - the solution should be much better

In your opinion, how should insurers' compliance with the moratorium on using genetic test results in life insurance be regulated?
[select all that apply]

☐ Self-regulation by the life insurance industry (FSC) [this is the current situation]
☐ Regulation through legally enforceable rules
☐ Other

Other:

__________________________________

In the UK, there is a moratorium that involves a formal agreement between the UK government and the Life Insurance Industry. Do you think a formal agreement between the Australian government and industry (Financial Services Council) is required on this issue in Australia?

◯ Yes
◯ No

[optional comment]

__________________________________
(Optional field)

Do you think the Australian government should introduce legislation to regulate the use of genetic test results in life insurance?

◯ Yes
◯ No

[optional comment]

__________________________________
(Optional field)

**As of 1 July 2019, please answer the following questions to the best of your knowledge**

| | True | False | I don't know |
|---|---|---|---|
| If a patient has an unfavourable genetic test result, their adult child must advise a life insurance company of the parent's genetic results when applying for a new insurance policy | ○ | ○ | ○ |
| Individuals with a current life insurance policy must notify their existing insurer if they get an unfavourable genetic test result | ○ | ○ | ○ |
| Life insurance companies are allowed to discriminate based on genetic test results, but health insurance companies are not | ○ | ○ | ○ |
| Life insurance companies have now agreed not to ask for genetic test results for policies worth up to $1 million | ○ | ○ | ○ |
| If a patient with an unfavourable genetic test result undertakes risk-reducing measures such as surveillance or surgery, an insurer must take this into account when assessing their risk | ○ | ○ | ○ |
| The moratorium also applies to travel insurance companies | ○ | ○ | ○ |

**ALMOST THERE!**

**Please indicate the degree to which you agree with the following statements:**

| | Strongly Agree | Agree | Disagree | Strongly disagree |
|---|---|---|---|---|
| The moratorium is easy to understand | ○ | ○ | ○ | ○ |
| The moratorium is easy to explain to patients | ○ | ○ | ○ | ○ |
| Patients are less confused than they used to be about insurance implications of genetic testing | ○ | ○ | ○ | ○ |
| Patients are more willing to have predictive genetic testing than they were before the moratorium was introduced | ○ | ○ | ○ | ○ |
| The moratorium has resolved some concerns I had about insurance discrimination | ○ | ○ | ○ | ○ |
| After the introduction of the moratorium, I still have concerns about insurance discrimination | ○ | ○ | ○ | ○ |
| Consumers are better protected post-moratorium than they were before the moratorium was introduced | ○ | ○ | ○ | ○ |

#### FINAL COMMENTS

In your opinion, what, if any, are/have been the benefits of the moratorium?

[optional]

(Optional field)

In your opinion, what, if any, are the limitations of the moratorium?

[optional]

(Optional field)

Do you have any final comments?

[optional]

(Optional field)

#### FURTHER CONTACT

As part of this research project, we may want to contact you to discuss the matters raised in this survey further. Any data collected in this follow-up interview will be de-identified before being published or shared.

If you consent to being contacted for a follow-up interview, please provide your contact details below.

◯ I prefer to remain anonymous
◯ I am happy to be contacted in the future

Name

__________________________________

Email address

__________________________________

Best telephone contact number

__________________________________
